# Analyzing Pain Patterns in the Emergency Department: Leveraging Clinical Text Deep Learning Models for Real-World Insights

**DOI:** 10.1101/2023.09.24.23296019

**Authors:** James A Hughes, Yutong Wu, Lee Jones, Clint Douglas, Nathan Brown, Sarah Hazelwood, Anna-Lisa Lyrstedt, Rajeev Jarugula, Kevin Chu, Anthony Nguyen

**Affiliations:** School of Nursing, Queensland University of Technology, Brisbane, Australia; Emergency and Trauma Centre, Royal Brisbane and Women’s Hospital, Brisbane, Australia; Australian e-Health Research Centre, CSIRO, Brisbane, Australia; QIMR-Berghoffer Research Institute, Brisbane, Australia; Metro North Health, Queensland, Australia; Faculty of Medicine, University of Queensland, Brisbane, Australia; Emergency Department, The Prince Charles Hospital, Queensland, Australia

**Author notes:** Corresponding Author: Dr James Hughes, Senior Lecturer, School of Nursing, Queensland University of Technology, N Block, Kelvin Grove Campus, Brisbane, Australia.

**Keywords:** Artificial Intelligence, Deep Learning Models, Electronic Health Records, Symptoms, Prevalence, COVID-19

## Abstract

**Objective:** To estimate the prevalence of patients presenting in pain to an inner-city emergency department (ED), describing this population, their treatment, and the effect of the COVID-19 pandemic.

**Materials and Methods:** We applied a clinical text deep learning model to the free text nursing assessments to identify the prevalence of pain on arrival to the ED. Using interrupted time series analysis, we examined the prevalence over three years. We describe this population pre- and post-pandemic in terms of their demographics, arrival patterns and treatment.

**Results:** 55.16% (95%CI 54.95% - 55.36%) of all patients presenting to this ED had pain on arrival. There were significant differences in demographics, arrival and departure patterns between those patients with and without pain. The COVID-19 pandemic initially precipitated a decrease followed by a sharp, sustained rise in the prevalence of pain on arrival, altering the population arriving in pain and their treatment.

**Discussion:** The application of a clinical text deep learning model has successfully identified the prevalence of pain on arrival. The description of this population and their treatment forms the basis of intervention to improve care for patients presenting with pain. The combination of the clinical text deep learning model and interrupted time series analysis has identified the effects of the COVID-19 pandemic on pain care in the ED.

**Conclusion:** A clinical text deep learning model has led to identifying the prevalence of pain on arrival and was able to identify the effect a major pandemic had on pain care in this ED.

## BACKGROUND

Historically, the quality of pain care in the emergency department (ED) has been unsatisfactory (1, 2). Although improvements have been made to pain treatments over the last 30 years, many fundamental aspects of pain care, such as the documentation of pain intensity on arrival and pain reassessment, need to be addressed (3). Inadequate documentation about pain and pain intensity can have immediate consequences for the quality of clinical care, and ongoing consequences for research and quality improvement because it makes it difficult to identify patients who have presented in pain. Instead, researchers typically rely on indirect measures, such as diagnosis or the administration of analgesic medication, to identify their target cohort when auditing medical records (4). However, such indirect measures will likely introduce bias into any resulting samples. The problem of how to accurately identify patients in pain has been brought further into the light with the widespread adoption of electronic health records (EHR) and the subsequent availability of “big data” to pain researchers. However, big data-level studies about ED presentations and pain care provided cannot be realized without changes to documentation practices, or the development of tools that provide easily reproducible and scalable ways to accurately identify patients arriving to the ED in pain.

Patients arriving to the ED with pain can be identified in audits of medical records by skilled clinicians reviewing information in the initial clinical assessment, even when pain severity is not documented (4, 6). However, the labor-intensive manual abstraction that is required limits the amount of data that can be reviewed. To address this issue we conducted two pilot studies in which human clinical abstractors were replaced by an “artificial intelligence” (AI) model using Natural Language Processing (NLP) and either Machine or Deep Learning. These conventional Machine Learning and Deep Learning models identified patients with pain from the triage (free text) nursing assessment with accuracies between 88% and 91% (7, 8). However, these studies have significant limitations regarding the adequacy of volume and quality of the training data available. The development of better training datasets (4) has made the development of better models possible. In testing, the model with the best performance has been a fine-tuned domain specific transformer-based deep learning model (9). This model was trained on a subset of data from the same hospital over the same period of time as used in this study. The model will be applied to a dataset of all presentations to a large inner city publicly funded ED over a three-year period with an aim to identify the prevalence of pain on arrival, and to describe this population. The COVID-19 pandemic occurred during the middle of data collection and provides a natural point to compare the effect (if any) of the pandemic on pain care in the ED.

### Objectives

1. Estimate the prevalence of pain on arrival over three years in a large inner-city ED.
2. Describe the population presenting in pain in terms of their demographics, treatment and outcomes compared to patients who did not present in pain.
3. Assess the impact of the COVID-19 pandemic on the prevalence of pain on arrival to the ED, the population presenting in pain and its treatment.

## MATERIALS AND METHODS

### Study Design

This is a cross-sectional study using data from two electronic health records that have undergone linkage. This study will be reported in line with the REporting of studies Conducted using Observational Routinely-collected health Data (RECORD) Statement (10).

### Study Setting

The Emergency and Trauma Centre (ETC) of the Royal Brisbane and Women’s Hospital (RBWH) is a principal referral and trauma centre for the state of Queensland in Australia. This ED sees approximately 85 000 primary presentations per year catering for adults requiring all specialties. The ED and hospital are publicly funded within a universal health care system.

### Data Sources

Two data sources were utilized for this study. The Emergency Department Information System (EDIS™) contains the patient presentation, demographic, assessment, diagnosis and deposition information. This includes the free text of the nursing triage assessment relied on for the classification task. The electronic medication dispensing system (PYXIS™) provided information on the pharmacological treatment of pain in the ED. The information contained within these systems was linked based on the hospital’s unique identifiers, admission and discharge dates and times. Records in either system where unique identifiers were missing were removed before linkage.

### Study Population

The study population included all patients who presented to the ETC of RBWH from 1^st^ March 2018 until 28^th^ February 2021. Patients who were under 18 years at the time of presentation (as this study focuses on adult patients in an adult facility), arrived from another hospital (and may have had significant treatment before arrival), or were missing unique identifiers (not allowing for data linkage) were excluded from the population. Figure 1 identifies the sample population and exclusions.

**Figure 1:**
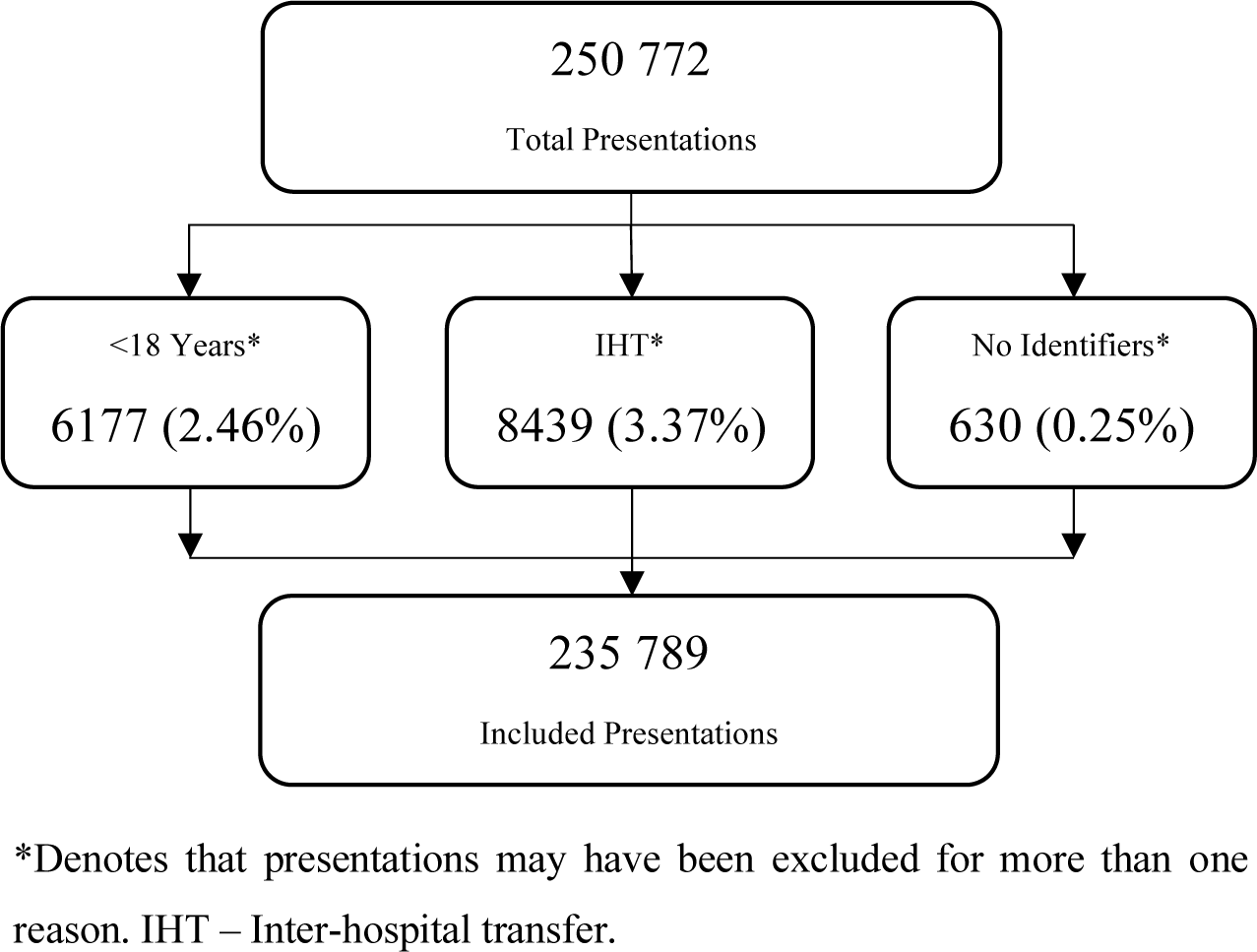
Study Population Flowchart

### Variables Collected

Previous applications of Symptom Management Theory to the study of pain in the ED guided the variables collected for this work (6, 11). Patients’ demographics, including socioeconomic status based on postcode, the workload of the ED on arrival, pharmacological treatment provided and health service outcomes such as time to first analgesic medication, were collected. A previous description of each variable collected has been published (4). These variables are summarized in Table 1. In addition to these variables, narrative triage nursing assessment was collected for the purposes of binary classification (pain/no-pain) by the clinical text deep learning model.

**Table 1:** Description of the population and differences between those presenting with and without pain.

| Variable |  | Total | Classification |  | Adjusted Odds Ratio<br>(aOR, 95%CI, p) |
| --- | --- | --- | --- | --- | --- |
|  |  |  | Pain | No-Pain |  |
| Demographics |  |  |  |  |  |
| Age (median, IQR) |  | 39.9 (27.7 – 57.6) yrs | 37.7 (26.9 – 54.6) yrs | 42.5 (28.8 – 61.5) yrs* | 0.996 (0.995 – 0.997, <0.001) |
| Sex (n, %) | Female | 117976 (50.0%) | 67029 (51.5%) | 54768 (51.8%)* | Ref. |
|  | Male | 117775 (50.0%) | 63007 (48.5%) | 50947 (48.2%) | 0.936 (0.919 – 0.953, <0.001) |
| IRSAD (median, IQR) |  | 1076 (IQR 1017 – 1096) | 1076 (IQR 1020 – 1096) | 1063 (1013 – 1096)* | 1.001 (1.001 – 1.001, <0.001) |
| Employment Status (n, %) | Employed | 92122 (39.1%) | 60036 (46.2%) | 32086 (30.3%)* | Ref. |
|  | Unemployed | 23565 (10.0%) | 9586 (7.4%) | 13979 (13.2%)* | 0.546 (0.527 – 0.565, <0.001) |
|  | Student | 14415 (6.1%) | 9519 (7.3%) | 4896 (4.6%)* | 0.859 (0.820 – 0.901, <0.001) |
|  | Pensioner | 41614 (17.7%) | 18578 (14.3%) | 23036 (21.8%)* | 0.655 (0.625 – 0.666, <0.001) |
|  | Other / Not Stated / | 64056 (27.2%) | 32330 (24.9%) | 31726 (30.0%)* | 0.707 (0.690 – 0.724, <0.001) |
|  | Unknown |  |  |  |  |
| Country of Birth (n, %) | Australia | 165681 (70.3%) | 88781 (68.3%) | 76900 (72.7%)* |  |
|  | Not – Australia | 70108 (29.7%) | 41270 (31.7%) | 28838 (27.3%)* |  |
| ATSI (n, %) | Indigenous | 11013 (4.7%) | 5523 (4.2%) | 5490 (5.2%)* | 1.065 (1.019 – 1.114, 0.006) |
|  | Non-Indigenous | 223065 (94.6%) | 123740 (95.2%) | 99325 (94.0%)* | Ref. |
|  | Unknown | 1651 (0.7%) | 751 (0.6%) | 900 (0.9%)* | 0.830 (0.731 – 0.942, 0.004) |
| RACF (n, %) | No | 231729 (98.3%) | 128406 (98.7%) | 103323 (97.7%)* | Ref. |
|  | Yes | 4060 (1.7%) | 1645 (1.3%) | 2415 (2.3%)* | 0.621 (0.578 – 0.665, <0.001) |
| Arrival |  |  |  |  |  |
| Mode of Arrival | Ambulance | 104693 (44.4%) | 52804 (40.6%) | 51889 (49.1%)* | Ref. |
|  | Walk-in | 125775 (53.3%) | 76607 (58.9%) | 49168 (46.5%)* | 1.085 (1.062 – 1.107, <0.001) |
|  | Other | 5321 (2.3%) | 640 (0.5%) | 4681 (4.4%)* | 0.215 (0.195 - 0.236, <0.001) |
| Day of Week | Monday | 35557 (15.1%) | 19697 (15.1%) | 15860 (15.0%)* |  |
|  | Tuesday | 33376 (14.2%) | 18321 (14.1%) | 15055 (14.2%)* |  |
|  | Wednesday | 33165 (14.1%) | 18258 (14.0%) | 14907 (14.1%)* |  |
|  | Thursday | 33208 (14.1%) | 17940 (13.8%) | 15268 (14.4%)* |  |
|  | Friday | 34072 (14.5%) | 18139 (13.9%) | 15933 (15.1%)* |  |
|  | Saturday | 32683 (13.9%) | 18484 (14.2%) | 14199 (13.4%)* |  |
|  | Sunday | 33782 (14.3%) | 19212 (14.8%) | 14516 (13.7%)* |  |
| Time of Day | Morning (0700 – 1459) | 102159 (43.3%) | 55552 (42.7%) | 46607 (44.1%)* | Ref. |
|  | Afternoon (1500 – 2259) | 94152 (39.9%) | 52564 (40.4%) | 41588 (39.3%)* | 1.016 (0.996 – 1.037, 0.123) |
|  | Night (2300 – 0659) | 39478 (16.7%) | 21935 (16.9%) | 17543 (16.6%)* | 1.069 (1.040 – 1.098, <0.001) |
| ATS | One | 2271 (1.0%) | 658 (0.5%) | 1613 (1.5%)* | Ref. |
|  | Two | 40462 (17.2%) | 18473 (14.2%) | 21989 (20.8%)* | 2.705 (4.426 – 3.017, <0.001) |
|  | Three | 85761 (36.4%) | 46752 (35.9%) | 39009 (36.9%)* | 6.452 (5.747 – 7.242, <0.001) |
|  | Four | 8225 (34.9%) | 52972 (40.7%) | 29278 (27.7%)* | 6.865 (6.113 – 7.711, <0.001) |
|  | Five | 25045 (10.6%) | 11196 (8.6%) | 13849 (13.1%)* | 2.183 (1.937 – 2.460, <0.001) |
| Stream | Acute | 148226 (62.8%) | 71052 (54.6%) | 77174 (73.0%)* | Ref. |
|  | Fast Track | 57517 (24.4%) | 41566 (32.0%) | 15951 (15.1%)* | 2.921 (2.844 – 3.000, <0.001) |
|  | Respiratory Isolation | 4852 (2.1%) | 2523 (1.9%) | 2329 (2.2%)* | 1.318 (1.238 – 1.403, <0.001) |
|  | Resuscitation | 25194 (10.7%) | 14910 (11.5%) | 10284 (9.7%)* | 4.271 (4.078 – 4.474, <0.001) |
| Departure |  |  |  |  |  |
| Departure Status | Admitted | 57410 (24.3%) | 20444 (15.7%) | 36966 (35.0%)* | Ref. |
|  | Short Stay Unit | 32215 (13.7%) | 21870 (16.8%) | 10345 (9.8%)* | 4.090 (3.956 – 4.429, <0.001) |
|  | DNW/LAMA/Unacceptable behavior | 12827 (5.4%) | 6247 (4.8%) | 6580 (6.2%)* | 2.134 (2.037 – 2.236, <0.001) |
|  | Death | 128 (0.1%) | 22 (0.0%) | 106 (0.1%)* | 0.505 (0.300 – 0.850, 0.010) |
|  | Discharged | 132401 (56.2%) | 80950 (62.2%) | 51451 (48.7%)* | 2.627 (2.558 – 2.698, <0.001) |
|  | Transfer | 808 (0.3%) | 518 (0.4%) | 290 (0.3%)* | 2.816 (2.406 – 3.296, <0.001) |
| 72 Hour Representation | Yes | 9834 (4.2%) | 4375 (3.4%) | 5459 (5.2%)* | 0.728 (0.695 – 0.763, <0.001) |
|  | No | 225955 (95.8%) | 125676 (96.6%) | 100279 (94.8%)* | Ref. |
| EDLOS (median, IQR) |  | 185 (112 – 291) min | 187 (120 – 281) min | 182 (98 – 307) min* | 1.001 (1.001 – 1.001, <0.001) |

IQR = Interquartile range, CI = Confidence Interval, yrs = Years, Ref = Reference Category, RACF = Residential Aged Care Facility, ATS = Australasian Triage Score, DNW = Did not wait, LAMA = Left Against Medical Advice, EDLOS = Emergency Department Length of Stay, min = Minute, IRSAD = Index of relative socioeconomic advantage and disadvantage; ATSI = Aboriginal or Torres Strait Islander.

### Classification of Pain/No Pain

A fine-tuned domain specific BlueBERT transformer-based deep learning model trained on 7000 manually coded presentations was used for this study (9). BlueBERT is a domain-specific pre-trained model that utilizes large-scale biomedical literature (PubMed abstracts) and clinical notes, and was fine-tuned to allow the large BlueBERT pretrained model to be trained specifically on the gold standard dataset (12). The gold standard dataset used for training, development and testing was created by manual abstraction of 10 000 presentations to the study setting by three experienced triage clinicians. These clinicians showed excellent agreement in their classification of pain/no pain based on the triage nursing assessment (κ = 0.807 (95% CI 0.795, 0.818), z = 80.703, p <0.001) (4). Previous work based on this dataset evaluated several classification models based on Deep Learning architectures, ranging from simpler Deep Learning architectures such as attention-based Recurrent Neural Network models to more complex general and domain-specific pre-trained transformer-based architectures. Through a learning curve analysis, a fine-tuned domain specific BlueBERT model trained on 7000 presentations was identified as having high accuracy (F_1_=0.9316) when tested on an evaluation dataset of 2000 presentations, with no further statistically significant increases in performances seen with larger training datasets (9, 13). This model was then deployed on the population in this study to undertake the classification task.

### Data Analysis

Data analysis was conducted in IBM SPSS Statistics version 28 (IBM Corp) and R version 4.2.2 (14)

#### Missing Data

With the exception of records missing unique identifiers or triage assessment no other records were removed because of missing data. All variables with missing data were reported on but remain in the descriptive and bivariate analysis. Cases with missing data underwent case-wise deletion in multivariable models.

Description of prevalence and the population

Prevalence (and its 95% confidence interval) is reported as a proportion of the population that is identified as arriving to the ED with pain. The population is described in terms of its demographics, arrival patterns, and departure patterns. Continuous variables are assessed for normality visually using P-P plots and described using means, standard deviations and 95% confidence intervals (or their non-parametric equivalents), categorical variables with counts and proportions. Differences in the populations presenting with and without pain were assessed using t-test (or Mann Whitney U test) and the chi-squared test of independence with statistical significance set at p<0.05. Variables where there is a significant difference between the pain and no pain group are entered into a multivariable logistic regression, where a variable remains significant (p<0.05) in the regression it is presented as an adjusted odds ratio. Differences in time to first analgesia between groups is assessed using the log-rank test as part of Kaplin- Meier analysis.

### Impact of COVID-19

The impact of the COVID-19 pandemic on the prevalence of pain on arrival to the ED is assessed using weekly timepoints in an Interrupted Time Series (ITS) Analysis via the its.analysis package (version 1.6.0) in R (15). Visual representations of the ITS analysis and diagnostics are prepared using the timetk package (16). The World Health Organization declared COVID-19 a pandemic on the 11^th^ March 2020 (17) and this date is used as the changeover point for the ITS analysis. The pre and post COVID-19 population presenting in pain are described using the same variables and the same statistical analysis as the population presenting in pain.

## RESULTS

### Missing Data

The age of the patient was not recorded in 448 (0.19%) records, sex in 38 (0.02%), postcode (to calculate the Index of Relative Socioeconomic Advantage and Disadvantage (IRSAD) score) in 14 958 (6.34%), employment status in 17 (0.01%), Indigenous (ATSI) status in 60 (0.03%). Significant amounts of missing IRSAD data have been reported in similar populations and has been shown to not be random with significant association to homelessness and international visitors/students (6). Therefore, no further analysis or imputation will occur.

### Prevalence of pain on arrival, description of the population and treatment received

The prevalence of pain on arrival to the RBWH ETC over a three-year period was 55.16% (95% CI 54.95% - 55.36%). Table 1 describes the population and compares the population presenting with and without pain on arrival.

The population presenting in pain were different to those not presenting in pain in terms of demographics, arrival patterns, urgency, treatment and deposition (Table 1). Overall, the person presenting in pain was less likely to be male, or have a higher socioeconomic status. The Indigenous Population were more likely to present in pain. Those from residential aged care facilities were less likely to present in pain. Patients arriving in pain stayed longer in the ED but were more likely to go home and not represent.

Pharmacological therapy remains the main treatment for pain in the ED. Of the patients identified as arriving in pain, 45.9% were identified as having an analgesic medication whilst in the ED with a median time to analgesia of 66 (IQR 38 – 117) minutes. Opioids remain the most common form of pain relief in the ED with 65.5% of all patients receiving pharmacological analgesia, receiving an opioid. There were 146 693 doses of analgesic medication delivered to the 59 667 patients in pain who received pharmacological treatment, representing 2.45 (range 0-22) doses per patient. The top analgesic medications dispensed are summarized in Supplementary Table 1. Patients presented with pain as a symptom of numerous conditions. The top 10 ICD-10 diagnosis for patients presenting in pain are described in Supplementary Table 2.

### Impact of the COVID-19 pandemic on the prevalence of pain on arrival

The following figure is an interrupted time series of the weekly prevalence of pain on arrival from March 2018 to February 2021 with separation of the pre and post COVID-19 pandemic periods.

The mean pre-pandemic incidence was 546 (SD 14.7) / 1000 presentations and the post-pandemic incidence was 562.9 (SD 18.9) / 1000 presentations. This equates to a mean difference of 16.9 patients presenting in pain per 1000 presentations (F(1,154)=18.710, p<0.001). Figure 2 demonstrates a level and slope change associated with the declaration of the pandemic. An initial decrease of approximately 25 / 1000 presentations was seen, which recovered to pre pandemic levels within nine weeks. This growth in incidence continues for 31 weeks prior to leveling off higher than the pre pandemic levels. Table 2 explores the differences in the populations presenting with pain pre and post pandemic.

**Figure 2:**
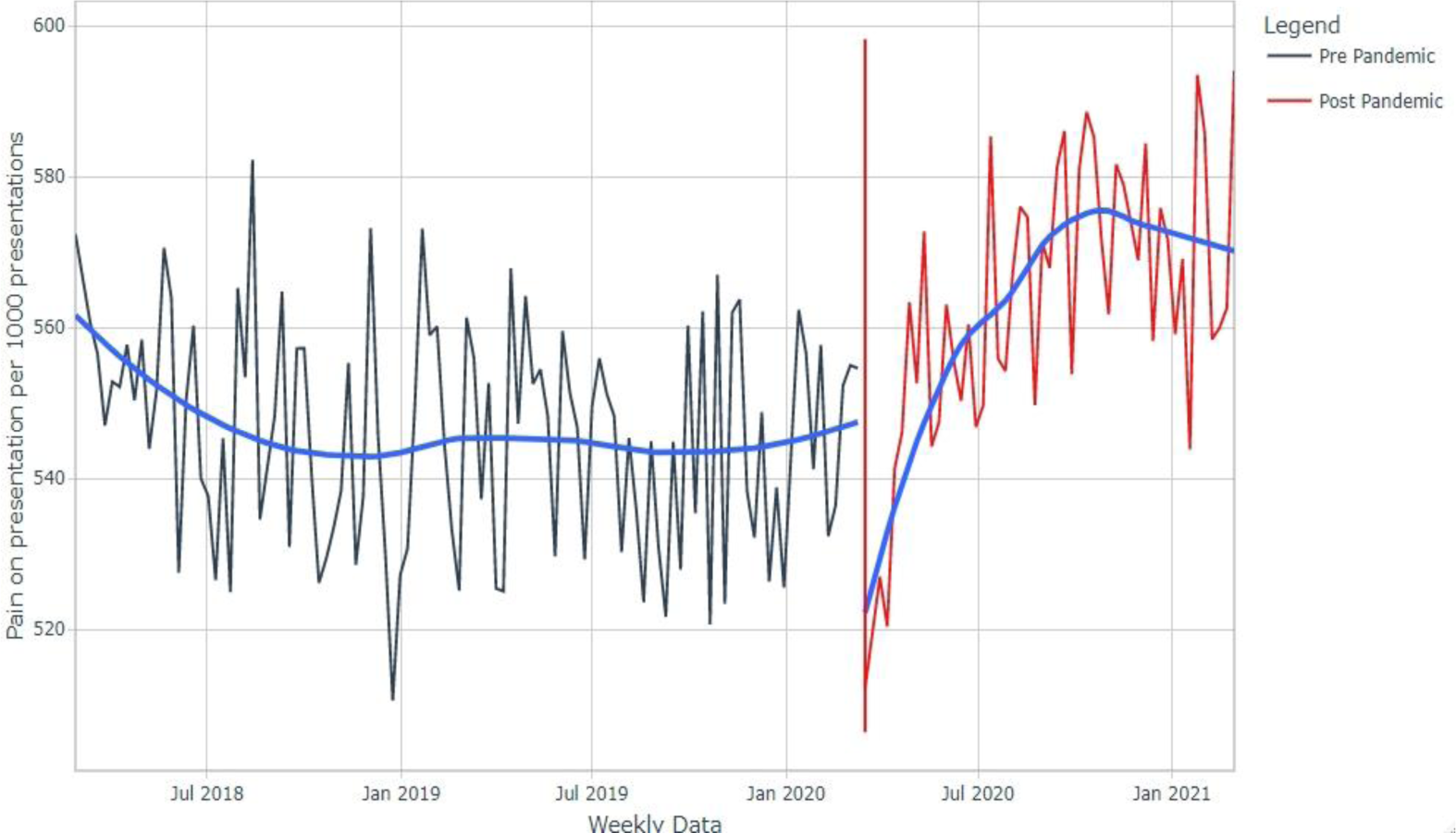
Interrupted time series plot of the rate of pain on presentation to the RBWH ETC pre and post the COVID-19 pandemic.

**Table 2:**
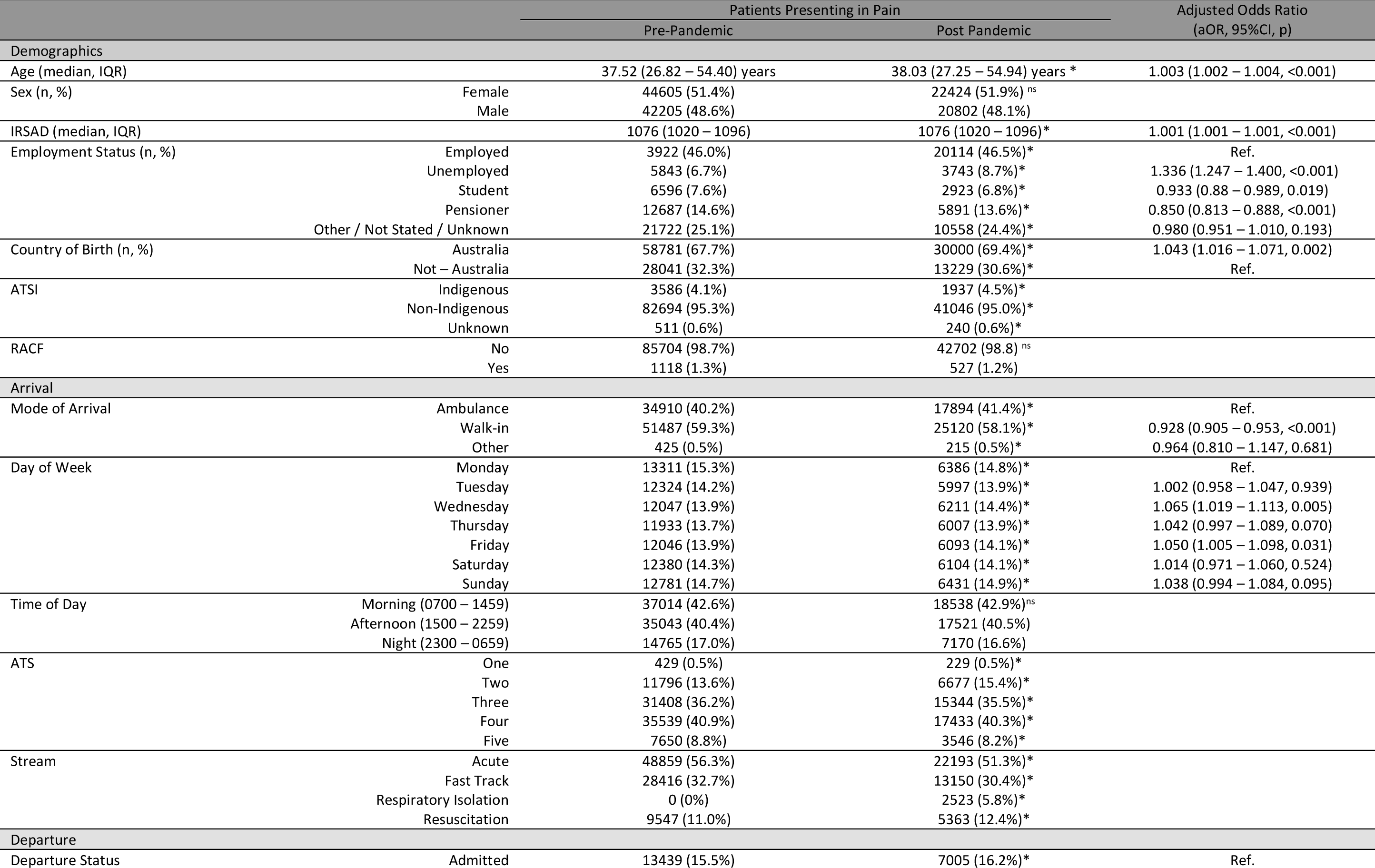
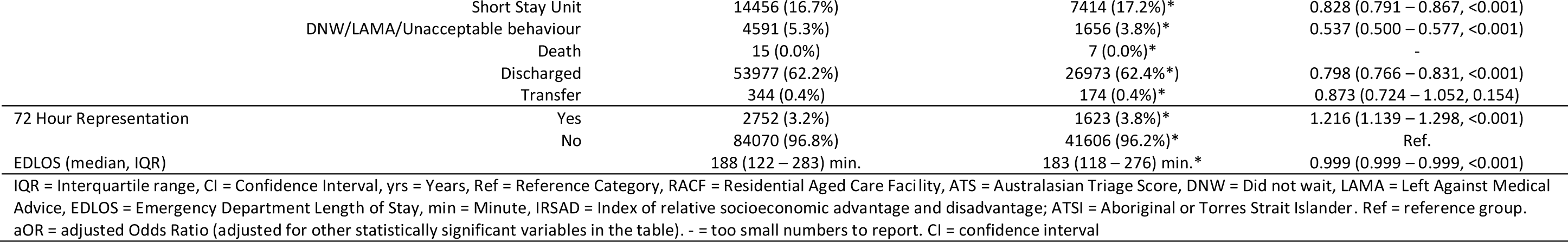
Differences in Demographics, Arrival and Departure metrics in patients presenting in pain pre and post pandemic.

The population presenting in pain post pandemic differed to the pre pandemic population in a number of key, interrelated factors. Patients post pandemic appear to be more unwell, as they were less likely to make their own way to the ED (aOR 0.928, 95%CI 0.905-0.953, p<0.001) and were significantly less likely to be discharged home (aOR 0.798, 95%CI 0.766-0.831, p<0.001) or to the short stay unit (aOR 0.828, 95%CI 0.791-0.867, p<0.001). Patients presenting in pain post pandemic were more likely to be unemployed (aOR 1.336, 95%CI 1.247-1.400, p<0.001) and born in Australia (aOR1.043, 95%CI 1.016-1.071, p=0.002). Despite a small decrease in the ED LOS (5 minutes), post pandemic patients were more likely to represent for further care a(OR 1.216, 95%CI 1.139-1.298, p<0.001).

In the pre pandemic period 52.17% of patients received analgesia and only 33.24% of patients received analgesia in the post pandemic period (difference 18.92% (95%CI 18.37% – 19.48%, p<0.001) (see Figure 3B). A similar reduction in the proportion of patients receiving opioids was observed (33.60% vs 22.87%, difference 10.73% (95%CI 10.23% - 11.24%), p<0.001) (see Figure 3C). The median time to analgesia (TTA) pre pandemic was 66 min (IQR 39 – 114) minutes and post 67 (IQR 37 – 127) minutes (p<0.001) which represents no clinically significant difference. However, when we consider the impact of the significant reduction in the proportion of patients receiving analgesia and the slightly longer TTA in Kaplan-Meier analysis, the differences become stark. There was a significant difference in the cumulative survival between the two groups (log-rank test ᵡ^2^ 4056.22(1df), p<0.001) (Figure 4).

**Figure 3:**
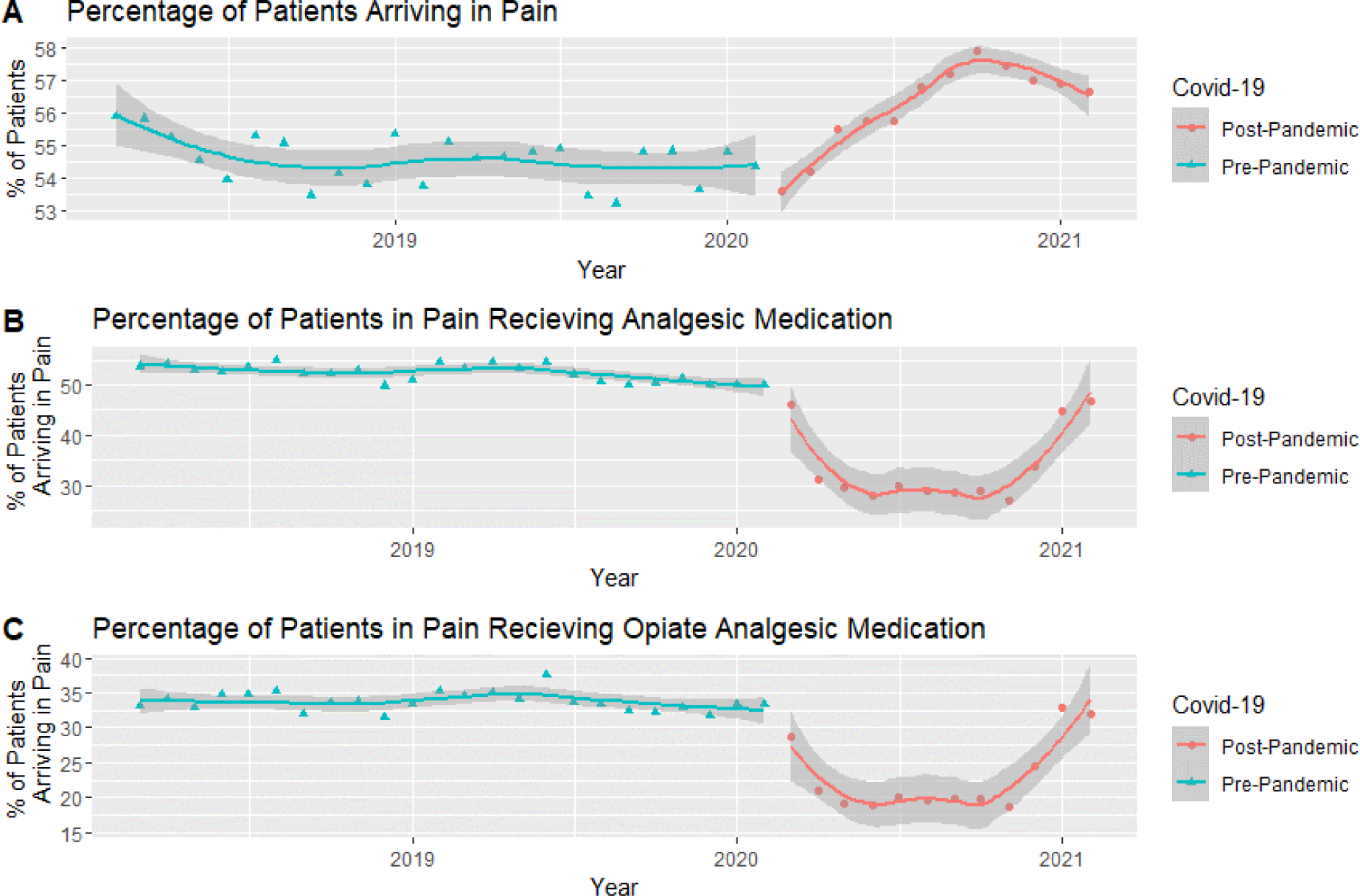
Proportions of patients A. Presenting in pain, B Receiving pharmacologic analgesia and C Receiving opiate analgesia per month pre and post the Covid-19 pandemic.

**Figure 4:**
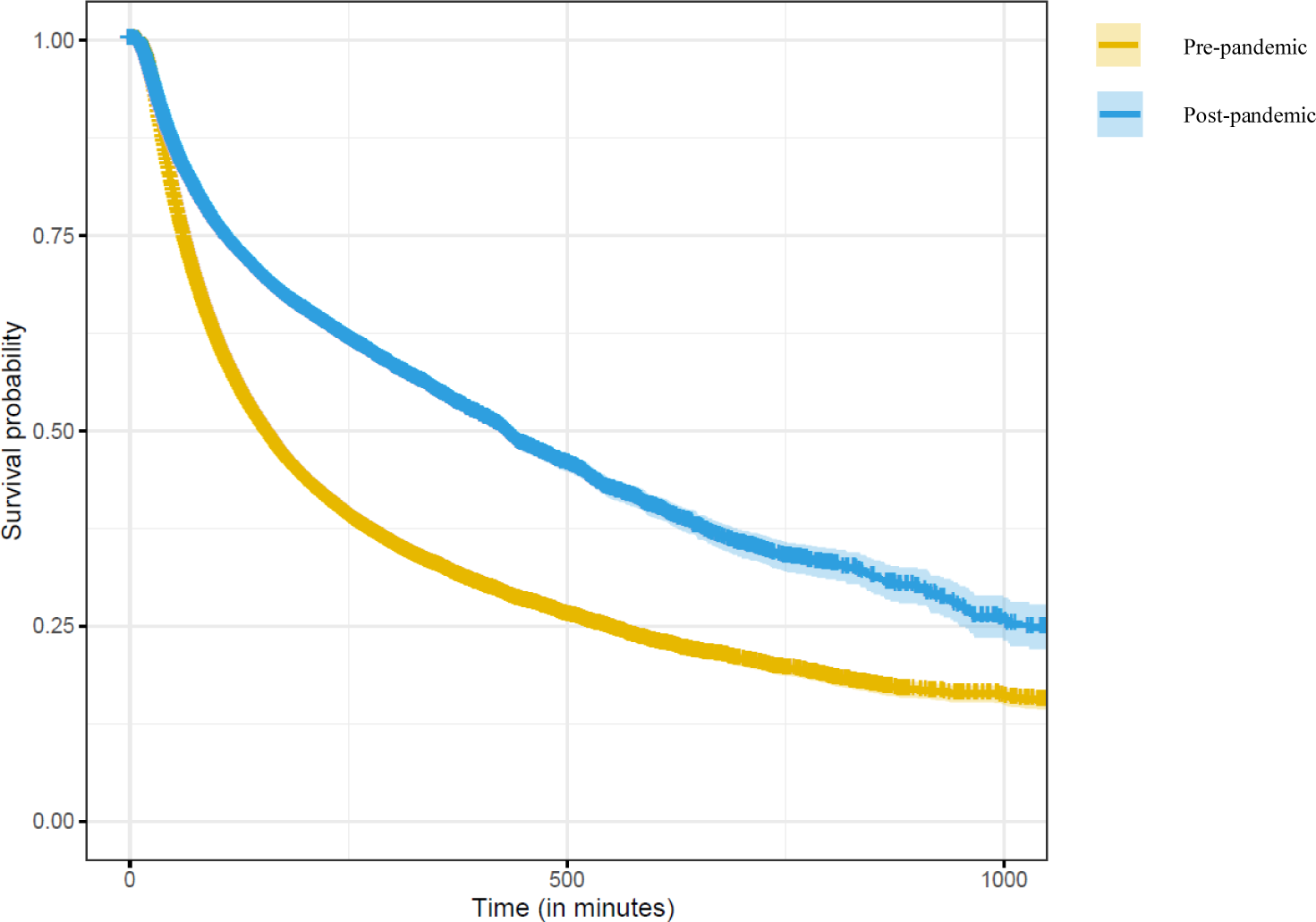
Survival Curve of the differences in time to analgesia pre- and post-pandemic with associated 95% confidence interval.

Post pandemic there was an increase in the use of oxycodone (7%), fentanyl (6%) and ketamine (1%) as can be seen in Supplementary Table 1. There was little change to the pre and post-pandemic ICD-10 diagnosis (Supplementary Table 2), with the addition of “Suspected Viral Illness” in the post pandemic period.

## DISCUSSION

A clinical text deep learning model (9) was successfully deployed on three years of data from one large inner-city adult ED. The outcome of this deployment is that the prevalence of pain on arrival was identified as 55.16% of all presentations to the ED, with a significant difference between pre- and post-pandemic prevalence. Pain remains the most common symptom on presentation to the ED, with over 55% of all patients experiencing pain on arrival in this department (4). Fundamentally the patient arriving in pain to the ED is different in terms of age, sex, socioeconomic status, ethnicity, residence, and urgency to those not in pain. However, the COVID-19 pandemic changed both the prevalence of pain on arrival and the population presenting in pain, demonstrating a shift in the reasons the community accessed this ED during the pandemic.

This study represents the application of a clinical text deep learning model in answering a clinical question related to pain in the ED. This methodology has been in development for four years (4, 7–9) and stems from other reports of the application of natural language processing to symptom identification in electronic health records (18–20). Emergency Medicine, with a large volume of undifferentiated patients, is an ideal setting for the deployment of artificial intelligence approaches to ED management, prediction of medical conditions and symptoms, patient acuity, deposition, and pre-hospital management (21, 22). The algorithm used in this work achieved an average accuracy of 93.2% (9) when compared to a manually coded random sample from the same dataset as used in this work (4).

The prevalence of pain on arrival reported by this methodology (55.16%) compared well to the reported prevalence in a manually coded sample from the same time period and setting (55.22%) (4). Previous reports of prevalence using retrospective methods have identified figures between 49.3% (23) and 62.3% (24) in a variety of EDs from multiple countries. The benefit of this methodology over those reported in these studies is the reproducibility and lack of reliance on documentation of pain, diagnosis of pain, pain intensity measurement or limitations of identified wordsearch. Previous work done in the same ED as this study has identified that pain is poorly documented, with only 33.1% of all patients presenting in pain having a pain intensity score documented (3) therefore reliance on this metric to identify a population may miss up to 67% of the population.

The characteristics of the population who present with pain to the ED in this study are fundamentally different to those who do not have pain on arrival. The population presenting in pain are younger, have a greater proportion of females, are more likely to be employed. Patients in pain are more likely to walk-in without pre-hospital care, presenting in the afternoon or night. When seen and treated by a provider are more likely to be discharged home or receive ongoing treatment in the Short Stay Unit. Previous studies have identified that the population presenting in pain are younger than those without pain and more likely to be female (24–26).

The COVID-19 pandemic changed the prevalence of pain on arrival to the ED, with an initial decrease and then an increase in the prevalence of pain on arrival. This corresponded with a change in the population presenting in pain and the treatment provided to these patients. Although the pandemic was declared in March 2020 (17) Queensland had it first case of COVID-19 in January 2020 and declared a public health emergency on the 29^th^ January 2020 (27). Initial decreases in the prevalence of pain on arrival correspond to lockdowns ordered in Queensland commencing on the 2^nd^ April 2020 and extended on the 9^th^ April (28). Both domestic and international borders were also either tightly controlled or closed at this time, not fully reopening until January 2022. Some of the changes in the population presenting in pain can be attributed to these changes such as patients more likely to be born in Australia due to the border closures and a higher proportion being unemployed due to labor issues associated with lockdowns. Other characteristics such as increased acuity (demonstrated by a higher proportion using ambulance services, and patients more likely to be admitted) may be representative of a hesitancy of the public to attend EDs during a pandemic, with a significant reduction in all ED attendances experienced in this ED (29) during this time. Previous work by Del Mar et al. (2023) has outlined the functional and operational changes made to this ED during the pandemic (29). With an increasing focus on managing patients in respiratory isolation, expediting admission or discharge, there was a reduction in the number of patients presenting with pain treated with analgesics or opiates. Other metrics such as an increasing representation rate for patients presenting in pain post-pandemic would indicate a level of inadequacy of care. However, without associated patient-reported outcome measures for this period of time we are unable to elaborate further on this phenomenon.

### Limitations

In general, deep learning models in healthcare suffer from a number of problems and challenges. In general, these are data insufficiency, model interpretability, privacy and ethical issues and heterogeneity (30). This study goes somewhat to addressing all these concerns. Data insufficiently was both measured and overcome in the development of this algorithm. The incremental multiphase framework used for model optimization having been previously published (9) from a manually coded dataset of 10 000 taken from the same population as this study (4). Interpretability remains difficult with deep learning models, however in the development of this approach we have previously identified a mechanism for interpretability within the task (8). Privacy and ethical issues are addressed within the robust nature of research governance within the jurisdiction this study was conducted. Like other applications of Artificial Intelligence, incorporation of the algorithm described into EHR may have potential ethical issues in the future, which may be justified by a low-risk, high benefit argument (31) in this eventuality.

## Conclusion

The use of a clinical text deep learning model has been effective at identifying the prevalence of pain on arrival to the ED from the narrative assessment completed on arrival to the ED. The ability to identify pain at a population level has allowed a robust description of the population, its treatment and the impact of a pandemic on these outcomes. Changes in care associated with the pandemic were closely tied to changes in society and the response of the healthcare system to this major event.

## Significance

This work has outlined a methodology for the identification of the patient in pain on presentation to the ED that is not reliant on pain intensity scoring, analgesic administration, or diagnosis. This removes many of the limitations of previous methods. This method allows researchers to take full advantage of the mass of EHR data available to them and assess interventions and associations at a population level rather than from representative samples.

## Data Availability

Due to local regulations this data is not available

## Acknowledgements

Nil

## Supplementary Material

**Supplementary Table 1:** Analgesia medication administered to patients presenting in pain.

| All Analgesia Administered (Top Ten Drugs) |  |  | Analgesia Pre-Pandemic (Top Ten Drugs) |  |  | Analgesia Post-Pandemic (Top Ten Drugs) |  |  |
| --- | --- | --- | --- | --- | --- | --- | --- | --- |
| Drug | Total | % | Drug | Total | % | Drug | Total | % |
| Paracetamol | 43849 | 29.9 | Paracetamol | 35353 | 31.2 | Oxycodone | 9995 | 30.1 |
| Oxycodone | 34997 | 23.9 | Oxycodone | 26194 | 23.1 | Paracetamol | 9516 | 28.6 |
| Ibuprofen | 22251 | 15.2 | Ibuprofen | 18027 | 15.9 | Fentanyl | 5659 | 17.0 |
| Fentanyl | 16936 | 11.5 | Fentanyl | 12436 | 11.0 | Ibuprofen | 4641 | 14.0 |
| Morphine | 9900 | 6.7 | Morphine | 7727 | 6.8 | Morphine | 2874 | 8.7 |
| Aspirin | 3353 | 2.3 | Aspirin | 2491 | 2.2 | Aspirin | 968 | 2.9 |
| Pantprazole | 1907 | 1.3 | Hyoscine-N-Butylbromide | 1439 | 1.3 | Pantprazole | 576 | 1.7 |
| Hyoscine-N-Butylbromide | 1740 | 1.2 | Pantprazole | 1361 | 1.2 | Ketamine | 447 | 1.3 |
| Odansetron | 1294 | 0.9 | Odansetron | 1158 | 1.0 | Ketorolac | 349 | 1.1 |
| Ketorolac | 1219 | 0.8 | Ketorolac | 899 | 0.8 | Hyoscine-N-Butylbromide | 345 | 1.0 |

**Supplementary Table 2:** ICD-10 Diagnosis of patients presenting in pain.

| All Painful Diagnosis |  |  | Pre Pandemic Diagnosis |  |  | Post Pandemic Diagnosis |  |  |
| --- | --- | --- | --- | --- | --- | --- | --- | --- |
| ICD 10 Diagnosis | Number | % | ICD 10 Diagnosis | Number | % | ICD 10 Diagnosis | Number | % |
| I20.0 Possible Cardiac Chest Pain | 4706 | 3.6 | I20.0 Possible Cardiac Chest Pain | 3181 | 3.7 | I20.0 Possible Cardiac Chest Pain | 1525 | 3.5 |
| R07.3 Non Cardiac Chest Pain | 3663 | 2.8 | R10.3 Abdo Pain - Localised To Lower Abdo | 2490 | 2.9 | R07.3 Non Cardiac Chest Pain | 1364 | 3.2 |
| R10.3 Abdo Pain - Localised To Lower Abdo | 3637 | 2.8 | R07.3 Non Cardiac Chest Pain | 2299 | 2.9 | R10.3 Abdo Pain - Localised To Lower Abdo | 1147 | 2.7 |
| R10.0 Abdominal Pain - Acute | 2942 | 2.3 | R10.0 Abdominal Pain - Acute | 1927 | 2.6 | R10.0 Abdominal Pain - Acute | 1015 | 2.3 |
| R51 Headache | 2576 | 2.0 | R51 Headache | 1763 | 2.2 | R51 Headache | 813 | 1.9 |
| S93.40 Ankle Sprain / Strain | 2248 | 1.7 | S93.40 Ankle Sprain / Strain | 1558 | 2.0 | S93.40 Ankle Sprain / Strain | 690 | 1.6 |
| S00.9 Minor Head Injury | 1946 | 1.5 | S61.9 Lacerated Finger | 1292 | 1.8 | S00.9 Minor Head Injury | 672 | 1.6 |
| S61.9 Lacerated Finger | 1918 | 1.5 | S00.9 Minor Head Injury | 1274 | 1.5 | S61.9 Lacerated Finger | 626 | 1.4 |
| S33.7 Low Back Pain | 1777 | 1.4 | S33.7 Low Back Pain | 1174 | 1.5 | S33.7 Low Back Pain | 603 | 1.4 |
| N39.0 Urinary Tract Infection | 1466 | 1.1 | N39.0 Urinary Tract Infection | 998 | 1.1 | Z11.5 Suspected Viral Disease | 481 | 1.1 |

